# High diversity of *Escherichia coli* causing invasive disease in neonates in Malawi poses challenges for O-antigen based vaccine approach

**DOI:** 10.1101/2024.09.24.24314145

**Authors:** Oliver Pearse, Allan Zuza, Edith Tewesa, Patricia Siyabuh, Alice J Fraser, Jennifer Cornick, Kondwani Kawaza, Patrick Musicha, Nicholas R Thomson, Nicholas A Feasey, Eva Heinz

**Affiliations:** Department of Clinical Sciences, Liverpool School of Tropical Medicine, Liverpool, UK; Malawi-Liverpool-Wellcome Programme, Kamuzu University of Health Sciences, Blantyre, Malawi; Queen Elizabeth Central Hospital, Blantyre, Malawi; Department of Vector Biology, Liverpool School of Tropical Medicine, Liverpool, UK; University of Liverpool, Institute of Infection, Veterinary and Ecological Sciences, Liverpool, UK; Kamuzu University of Health Sciences, Malawi; Wellcome Sanger Institute, Parasites and Microbes Program, Hinxton, UK; London School of Tropical Medicine and Hygiene, Department of Pathogen Molecular Biology, London, UK; The School of Medicine, University of St. Andrews, St. Andrews, UK; University of Strathclyde, Strathclyde Institute for Pharmacy and Biomedical Sciences, Glasgow, UK

**Keywords:** Neonatal infection, neonatal sepsis, neonatal meningitis, vaccines, sero-epidemiology, O:H-type, H-antigen, antimicrobial resistance, sub-Saharan Africa

## Abstract

*Escherichia coli* is an important cause of neonatal sepsis and the third most prevalent cause of neonatal infection in sub-Saharan Africa, often with negative outcomes. Development of maternally administered vaccines is under consideration, but to provide adequate protection, an understanding of serotypes causing invasive disease in this population is essential. We describe the genomic characteristics of a collection of neonatal *E. coli* isolates from a tertiary hospital in Blantyre, Malawi, with specific reference to potential protection by vaccines under development. Neonatal blood or cerebrospinal fluid cultures from 2012-2021 identified 205 *E. coli* isolates, and 170 could be recovered for sequencing. There was very high diversity in sequence types, LPS O-antigen-type and fimbrial H-type, which all showed temporal fluctuations and previously undescribed diversity, including ten putative novel O-types. Vaccines in clinical trials target the O-antigen but would only protect against one third (33.7%) of neonatal sepsis cases in this population (EXPEC9V, in clinical trials). An O-antigen based vaccine would require 30 different O-types to protect against 80% of infections. Vaccines against neonatal sepsis in Africa are of considerable potential value, but their development requires larger studies to establish the diversity and stability over time of relevant O-types for this population.

**Data summary:** All sequencing data is freely available under the sequencing project IDs ERP120687 (short read data; accessions in Supplementary Table 1) and PRJNA1121524 (long-read data; accessions in Supplementary Table 2), detailed per-isolate information is provided in Supplementary Table 3. Blood culture and CSF data used to show the trends and numbers of *E. coli* cases per year is available in Supplementary Table 4.

## Introduction

Neonatal infection is the third largest cause of neonatal death worldwide, currently at 18 per 1,000 live births globally. Sustainable Development Goal 3.2^1^ aims to reduce this to 12 per 1,000 live births by 2030. *Escherichia coli* is the third most prevalent cause of neonatal infection in sub-Saharan Africa^2^ and is a particularly important cause of early onset sepsis^3^ (EoS; sepsis before 72 hours of life).

Antimicrobial resistant (AMR) *E. coli* infection makes this problem worse and is an increasing problem in countries in sub-Saharan Africa^4^, such as Malawi. Limited access to antibiotics in the countries with the highest burden of infection makes these infections even more lethal^5^. In Malawi for example there are few antimicrobial choices for neonates beyond the first-line (benzylpenicillin and gentamicin) and second-line (ceftriaxone) therapies, for which there is already resistance amongst *E. coli* isolates^6^. In this context, innovative approaches to preventing infection with *E. coli* are required. One proposed approach is maternal administration of vaccines to give neonates passive immunity as is already used for *Bordetella pertussis^7^*and being developed for Group B *Streptococcus*^8^. This would reduce the deaths and prolonged hospital stays of neonates linked to *E. coli* sepsis and meningitis, reduce the use of antimicrobials and thus the population-level spread of AMR. However, the surface exposed structures that could be targeted by vaccination have high levels of diversity. There are currently 186 known O-polysaccharide antigens (O-types) for *E. coli*, 67 known capsular antigens (K-types) and 53 known flagellar antigens (H-types)^9^. O-types and H-types are most important as they are expressed by all *E. coli* and recognized by the immune system of the host. Crucially, only a small proportion of these serogroups are thought to cause the majority of invasive disease^10^ but this can vary depending on location and patient characteristics. A vaccine would therefore need to target the correct surface exposed structures for the patient population in question.

*E. coli* vaccines targeting the O-antigen are in development, including the ExPEC4V^11^ and ExPEC9V^12, 13^ in clinical trials. These 4-valent and 9-valent vaccines have been produced to target the most prevalent serogroups causing invasive *E. coli* infection in older adults in high-income countries. Whether these candidates would reduce neonatal sepsis in sub-Saharan Africa is unknown, and addition of other types may be challenging; one O-antigen already had to be removed from the initially 10-valent ExPEC9V as the functional antibody assay for the O8 *E. coli* strain was not able to distinguish an immunological response to vaccination^13, 14, 15^.

To our knowledge there are no studies specifically examining the antigenic diversity in *E. coli* causing invasive infection in neonates in sub-Saharan Africa, a population for which an understanding of this diversity is crucial if vaccination is to be feasible and to achieve equity in coverage across different regions. This study presents a genomic description of an unbiased collection of neonatal *E. coli* isolates, collected from 2012 to 2021 from neonates in a Malawian hospital, with a focus on O- and H-type diversity.

## Methods

### Setting

Queen Elizabeth Central Hospital (QECH) is a government run tertiary referral hospital for the Southern Region of Malawi; care is free at the point of delivery. It directly serves urban Blantyre (population ∼800,000 as of 2018 census data). Chatinkha nursery receives approximately 5,000 admissions a year, with between 30 and 90 neonates on the ward at any one time. It admits neonates that have not gone home; either those that were born in QECH or those referred from another hospital. Paediatric nursery receives neonates that have been admitted from home via paediatric A & E. In both of these wards there is 24-hour nursing care, and daily medical ward rounds. There is access to diagnostic blood and cerebrospinal fluid (CSF) culture, continuous positive airway pressure, oxygen, IV fluids, blood transfusion, radiology, and blood testing. There is also access to an on-site HDU. The PICU/HDU and paediatric surgical ward are located on the Mercy James Hospital, which opened in 2017. This is a surgical hospital on the QECH site but it is in a separate building, managed separately and philanthropically funded. At the surgical hospital there is additionally access to intensive care facilities, which allows for the use of vasopressors and intubation.

### Microbiological sampling and processing

Routine, quality assured diagnostic blood culture services have been provided to the medical and paediatric wards by the Malawi-Liverpool-Wellcome Programme (MLW) since 1998. Briefly, 1-2mL of blood was taken from neonates (up to 28 days old) with risk factors for sepsis (i.e. maternal fever during labour, prolonged rupture of membranes, tachypnoea), or clinical suspicion of sepsis (fever >38°C, tachypnoea, tachycardia, reduced activity, seizures). For some clinical records, information on age was only described in months of age and individuals whose age was entered as being ‘1 month’ were also considered as neonates for the purposes of our study. For neonates with clinical suspicion of sepsis or other clinical suspicion of meningitis (raised fontanelle, abnormal neurology), a lumbar puncture was also performed.

Blood was collected using aseptic methods and inoculated into a single aerobic bottle (BacT/Alert, bioMérieux, Marcy-L’Etoile, France), then incubated using the automated BacT/Alert system. Samples that flagged positive were Gram stained and Gram-negative bacilli were identified by Analytical Profile Index (bioMérieux). Antimicrobial susceptibility testing was determined by the disc diffusion method (Oxoid, United Kingdom). All *E. coli* isolates were tested for their susceptibility to ampicillin, cefpodoxime (as an ESBL screen), chloramphenicol, ciprofloxacin, co-trimoxazole and gentamicin, and those that were resistant to cefpodoxime also had their sensitivity tested against amikacin, co-amoxiclav, cefoxitin, meropenem, pefloxacin and piperacillin-tazobactam. BSAC breakpoints were used until 2018, at which time EUCAST breakpoints were introduced. Details for the identification of other organisms are described elsewhere^4^. *E. coli* isolates were stored at -80°C on microbank beads.

Between 1998-2010, diagnostic results were entered into ledgers that were later digitized, and from 2010, PreLink, a Laboratory Information Management system was used and information stored on an SQL database. The MLW database was screened from 2000 to 2021 to identify all cases of *E. coli* infection in the hospital, and all *E. coli* isolates from neonates (recorded as less than 29 days old or as 1 month old on ledgers) in the period from September 2012 to March 2021 were selected for whole genome sequencing. This time period was chosen as this was the time period for which we had consistent metadata at the time of whole genome sequencing.

### Whole genome sequencing

A single microbank bead was removed from all selected *E. coli* isolates, which was thawed, streaked on MacConkey’s media and this media incubated for 18-24 hours at 37°C. Plates with growth of a single colony type then had a single colony pick taken and inoculated into 15ml of buffered peptone water for 18-24 hours at 37°C. These samples were then centrifuged and the supernatant was discarded. The pellet was then resuspended in buffer. For the short-read sequencing the DNA was extracted using the QIAsymphony machine and QIAsymphony DSP kit with onboard lysis, according to the manufacturer’s instructions. Quality control was done using Qubit and samples with a DNA volume of less than 200ng were repeated. Samples that passed QC underwent Whole Genome Sequencing (WGS) at the Wellcome Sanger Institute at 364 plex on the Novaseq SP generating 150bp paired-end reads.

For long-read sequencing of selected isolates, DNA was extracted using the MasterPure Complete DNA and RNA isolation kit following the manufacturer’s instructions for the purification of DNA from cell samples. DNA was then quality controlled using the Qubit dsDNA Broad Range assay and the TapeStation (4150) system, using the Genomic DNA Screen Tape Kit.

Long-read sequencing was performed on a MinION MK1B sequencing device (ONT, U.K.). Library preparation was carried out according to the manufacturers protocol, using the ligation sequencing kit (SQK-LSK109) and Native Barcoding Expansion Kits (EXP-NBD104; all ONT). Sequencing was carried out using a FLOW-MIN106 R9.4.1 flow cell (ONT). Two samples did not produce sufficient data and were re-sequenced using the Native Barcoding Kit 24 V14 (SQK-NBD114.24, ONT), following the manufacturer’s instructions. Sequencing was then carried out using a R10.4.1 Flongle flow cell (ONT).

### Genomic sequence analyses

Species confirmation was performed using Kraken v1.1.1^16^, and any sample with greater than 5% read content other than *E. coli* or Unclassified was excluded. Annotated assemblies for the short read data were produced using the pipeline described previously^17^. De novo assembly of genome sequences was performed using SPAdes v3.14.0^18^, trialing different kmer lengths between 41 and 127 to find the optimal kmer length. An assembly improvement step was applied to the assembly with the best N50 and contigs scaffolded using SSPACE v2.0^19^ and sequence gaps filled using GapFiller v1.11^20^. Assembly statistics were generated using the Sanger Pathogens pipeline as available on github^21^.Samples with <20 or >200 contigs and a genome size of <4.4MB or >5.6MB were excluded. Isolates with greater than 5% heterozygous SNPs of the total genome were also excluded due to potential within-species contamination. Automated annotation was performed using PROKKA v1.5^22^ and genus specific databases from RefSeq^23^. The improved assembly step uses software developed by the Pathogen Informatics team at the WSI which is freely available for download from GitHub^24^ under an open-source license, GNU GPL 3. The improvement step of the pipeline is also available as a standalone Perl module from CPAN^25^.

Sequence type (ST) was determined using mlst v2.23^26,27^. AMRFinderPlus v3.10.40 was used to identify AMR genes^28^. SRST2 v0.2.0^29^ with the EcOH database^30^ was used to determine the O- and H-types for the bacterial isolates. Where an isolate had more than one predicted O- or H-type, both sets were counted. We further aimed to confirm the subtypes (O1A, O6A, O18A, O25B) of relevance for the vaccine, which are not distinguished from their related (but immunologically non-identical) subtypes when using automated prediction via SRST2. For the distinction of O1A and O1B, sequences were compared to the specific primers and probe used previously^31^. For O18, there are four distinct subtypes described: O18A, O18A1, O18B and O18B1. We distinguished O18ac (∼O18A/A1) from O18ab (∼O18B/B1) using the presence/absence of an IS element inserted at a location immediately upstream of the *wzz* gene^32^. To distinguish O25A and O25B, we assessed the operon structure as described previously to distinguish these types^33^. We were not able to identify any description on the genetic differences of O6A and other O6 sub-types (referred to hereafter as O6?).

To further investigate their antigenic structure we performed long read sequencing using the Oxford Nanopore platform for 14 selected isolates. Basecalling and demultiplexing on raw long-reads was performed with guppy v 2.6.1^34^ using the super-accurate model for basecalling, adapters removed with porechop v 0.2.4^35^, and low-quality reads were removed with filtlong v0.2.2^36^ before assembly. Long-read-first hybrid assemblies from isolates sequenced on the Oxford Nanopore platform were produced using Flye v 2.9.3^37^, then visualised with Bandage v0.8.1^38^. Long-read polishing was performed using Medaka v1.8.0; https://github.com/nanoporetech/medaka^39^, then short-read polished with Polypolish v0.6.0^40^ and Pypolca v0.3.0^41^.

Assembled genomes were annotated using prokka as described above, and the O:H loci were investigated initially using the ECTyper^42^ which uses assemblies as input, which yielded comparable results to the srst2 EcOH search (see Supplementary Table 4); we then assessed these isolates further manually to determine the exact structure of these untyped, potentially novel O-Ag types.

### Statistical analysis

Statistical analysis was done in the R statistical programming language^43^, using Rstudio^44^ and the packages here^45^, tidyverse^46^ and lubridate^47^. Graphical data representation was performed using the packages ggplot2^48^, ghibli^49^, RColorBrewer^50^, pals^51^, MetBrewer^52^, gggenes^53^ and ggpubr^54^.

*E. coli* sample positivity was calculated as the number of samples which were positive for *E. coli* per 1,000 samples taken (blood culture or CSF) according to the equation below.

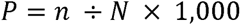

Where *P* is the sample positivity rate, *n* is the number of *E. coli* infections and *N* is the total number of blood culture or CSF samples taken.

The change in AMR over time was estimated by regressing resistance pattern of isolates against year of occurrence using a generalized linear model in R (glm function) with the binomial family and a logit link statement. Isolates were given a binary categorization of 1 if they were resistant or had intermediate resistance to an antibiotic and 0 if they were sensitive. Plots of AMR trends over time had lines of best fit that utilized a linear model.

For the rarefaction curves in the main text, lines showing hypothetical coverage for vaccines based on the *n* most frequent O-types or H-types were compared to lines showing the hypothetical coverage based on the EXPEC4V and EXPEC9V vaccine O-types; the supplementary data furthermore shows our analysis for the original ExPEC10V composition. For isolates with more than one allele for H-type, both were included in the rarefaction curve. For H-types the rarefaction curve goes above 1 as there were multiple isolates with more than one H-type. For O-types there was only one O-type per isolate (there was one O-type with both O8 and O160 sections, but it is unclear whether this expresses both sugar molecules or is a hybrid O-antigen) so the rarefaction curve only goes to 1. The analysis was repeated for the supplementary materials but only counting each isolate with multiple H-types once (the H-type that had the highest population frequency was chosen). The other allele was ignored, as theoretical protection from the vaccine was assumed by the H-type or O-type that was most prevalent.

### Ethics statement

This study was ethically approved by the Kamuzu University of Health Sciences College of Medicine Research Ethics Committee (COMREC P.06.20.3071). The ID numbers used in this manuscript are specimen IDs generated by the MLW diagnostic laboratory and not patient IDs; only the research team and members of the clinical staff in hospital that have access to the password protected laboratory information management system at MLW would be able to make the link to an individual.

## Results

### Clinical characteristics

There were 3394 *E. coli* isolated from 264692 blood culture and CSF tests over the period from 2000 - 2021. The number of cases of *E. coli* per year for all ages ranged from 88 to 233, with an average of 139 cases per year. The number of cases of *E. coli* per year for neonates ranged from 4 to 41, with an average of 15 cases per year. (Figure 1A). The number of blood culture or CSF samples taken per year for all ages ranged from 2796 to 26230 with an average of 11028.8 in a year. The number of blood culture or CSF samples taken per year for neonates ranged from 77 to 3211 with an average of 1227.7 per year. The positivity rate per 1,000 blood culture or CSF samples for all age groups was highest in 2006 and lowest in 2013, with an average positivity rate of 14.1/1000 samples/year. For neonates it was highest in 2004 and lowest in 2015, with a similar average positivity rate of 13.7/1,000 samples (Figure 1B).

**Figure 1A).**
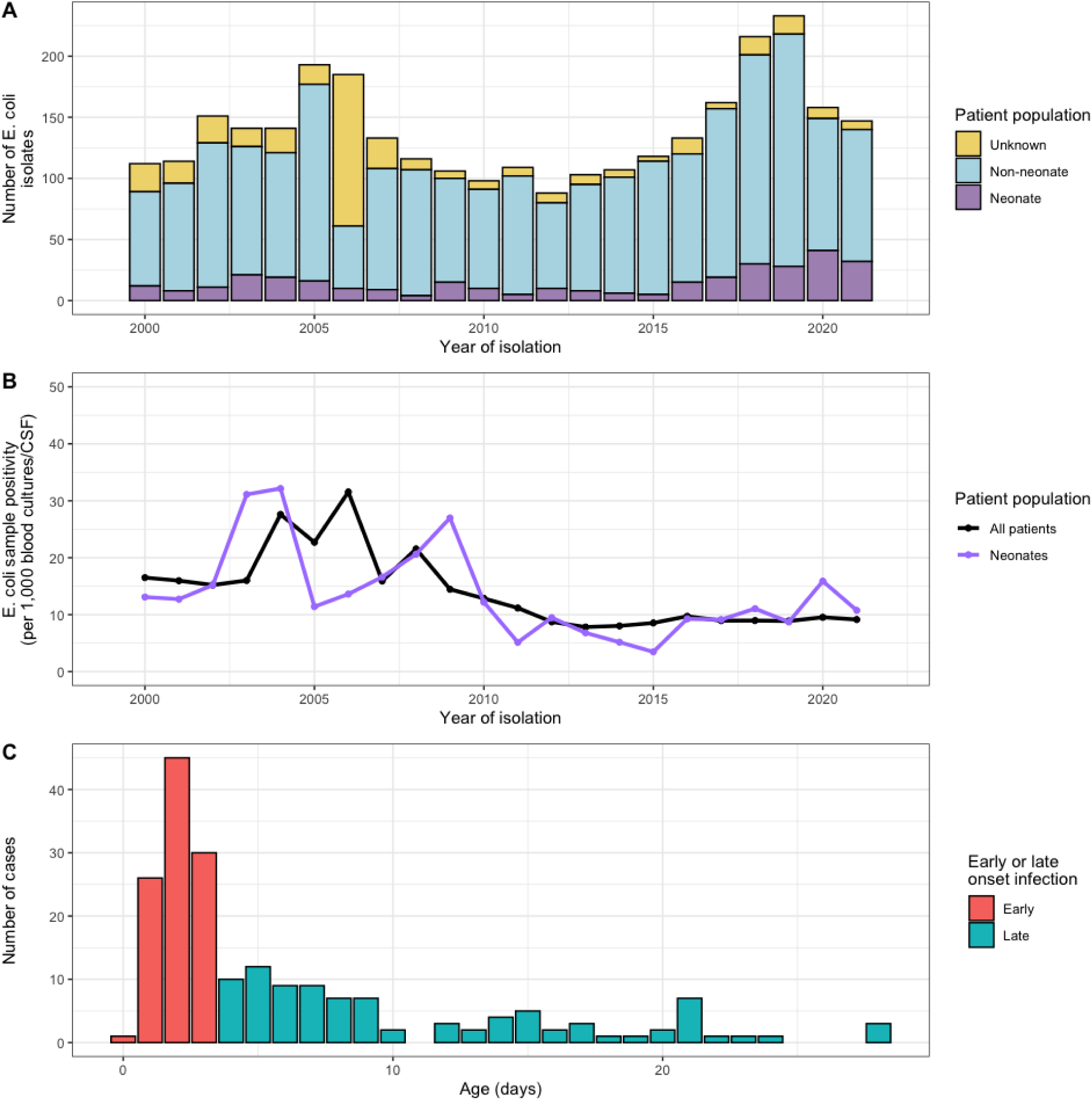
Numbers of E. coli cases per year at QECH. Bars represent the crude frequency of E. coli infection for each year from 2000 - 2021, with the different colours representing the different age groups of the patients. B) Blood culture and CSF positivity rate (per 1,000 blood culture or CSF samples) of E. coli in neonates and the entire patient population (including neonates). C) Age range of neonates in the current study, colours highlighting early (< 72 hours of life) or late (>72 hours of life) onset infection.

We identified 201 *E. coli* isolated from neonates in the period from September 2012 to March 2021 (Figure 2C); 95/201 (47.3%) were female, with a median age of 3 [IQR 2 - 8] days (Figure 1D). Early onset sepsis accounted for 109/201 (54.2%) of cases, with late onset sepsis accounting for the rest (Figure 1C). Of these isolates 163/201 (81.1%) were cultured from blood and 38/201 (18.9%) from CSF.

**Figure 2A).**
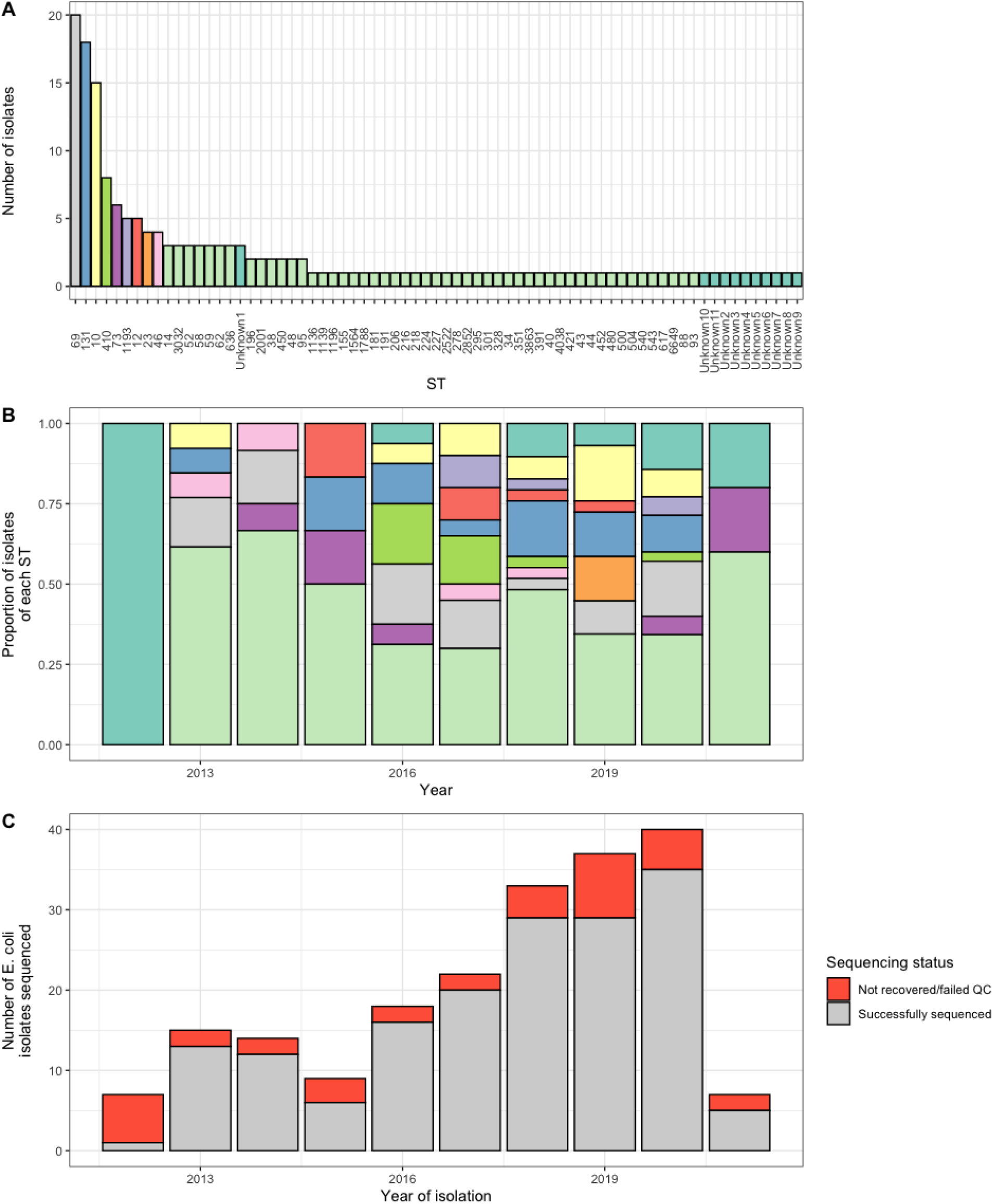
Frequency of different STs in the collection. B) Frequency of the different STs by year. C) Number of E. coli isolates selected for WGS for this current study. 2012 and 2021 were years for which isolates were only selected for part of the year.

There were 110/201 (54.7%) cases from the Chatinkha nursery, 62/201 (30.8%) from paediatric nursery, 12/201 (6%) were from the paediatric A&E, 12/201 (6%) were from the PICU/HDU and 110/201 (54.7%) were from the paediatric surgical ward (Supplementary Figure 1).

Of these 201 isolates, 192 were recovered for WGS and 170 passed QC (22 failed; Figure 2C). 17 isolates were excluded due to contamination, and 17 were excluded due to failure of assembly or poor assembly metrics (twelve isolates failed both, and five of each just failed due to one reason). There were no isolates that were excluded due to potential within-species contamination. There was one duplicate isolate (two sequencing runs of the same isolate from the same sample), of which one isolate was removed from analysis, so the total number of isolates analysed was 169.

### Population structure

There were 71 different STs represented in the collection (Figure 2A & B; 60 typed and 11 untyped). The most frequently isolated STs were ST69 with 20/169 (11.8%) isolates, ST131 with 18/169 (10.7%) isolates, ST10 with 15/169 (8.9%) isolates and ST410 with 8/169 (4.7%) isolates. There were 13/169 (7.7%) isolates with untyped STs using MLST which represented eleven different MLST allele patterns (ten singles and three of the same allele pattern). Over half of the observed STs were only represented by a single isolate (38/71, 53.5%) showing that neonates tested here were exposed to and infected by a highly diverse pool of *E. coli* that span the species phylogeny. ST410 was disproportionately found in the CSF rather than blood culture samples (6/8 [75%]), compared to ST69 (2/20 [10%]), ST131 (2/18 [11%]) and ST10 (2/14 [14%]) which were found primarily in blood culture samples.

Importantly, the ST diversity was also highly variable over time. 50/71 (70.4%) STs were only found in a single year, and 7/71 (9.9%) STs were found in only two years, with each year showing a similar pattern of high diversity during the entire study period. Frequently occurring STs were also prominent in different years (e.g. ST410 in 2016 and 2017), only ST69 was consistently isolated and was the most frequently isolated or joint most frequently isolated ST in 5 out of 7 of the years with more than ten isolates. Even the most prevalent STs like ST69 or ST131 however fluctuated over time, with numbers between 1/29 (3%) and 6/35 (17%) for ST69, and 1/20 (5%) and 5/29 (17%) for ST131, respectively, with no clear trend over time observable for any of the main STs. We also note that untyped isolates are derived from a range of years, including the most recent data. This indicates that these are not representing older lineages that might not be covered well in databases consisting mainly of recent samples, but indicating a high undescribed diversity circulating at present time. Two samples (one blood culture and one CSF) were polymicrobial. They both contained two different colony morphologies. The CSF sample contained one isolate of ST14 and one of ST69 and the bloodstream sample contained one isolate of ST1136 and one of ST10.

### O-antigen and H-antigen diversity

There were 63 O-types found in the collection, none of which were identified in more than 10% of the isolates. The most frequently isolated were O15 with 15/169 (8.9%) isolates, O25B with 15/169 (8.9%) isolates and O8 with 13/169 (7.7%) isolates (Figure 3A). These same O-types (O15, O25B and O8) were also the only ones found in greater than 75% of the years (6 out of 7 or more) that had more than 10 isolates (Figure 3B). Similar to ST types there was no sign of the population becoming increasingly dominated by any types over time, the composition between years differed strongly (Figure 2B). There were no years in which any O-type represented greater than 20% of the isolates, the largest proportion of isolates belonging to a single O-type per year were O11 and O8 which were both associated with 3/16 (18.8%) of all cases in 2016 (Figure 3B).

**Figure 3A).**
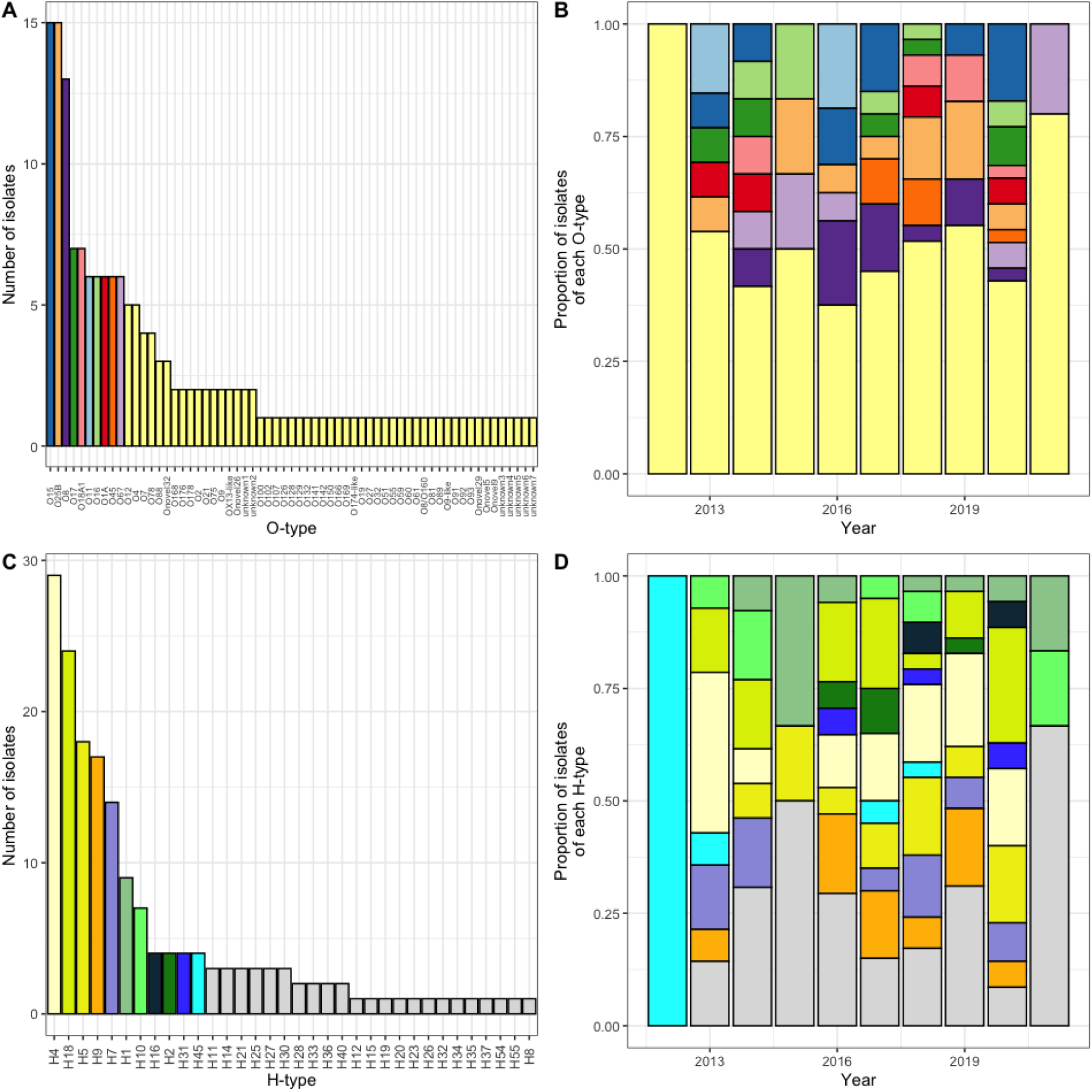
A bar chart showing the frequency of the different O-types. Where an isolate had more than one O-type gene, this was counted twice. B) A bar chart showing the proportion of isolates per year that had different O-types. The colours are the same as those represented in Figure 3A.C) A bar chart showing the frequency of the different H-types. Where an isolate had more than one H-type gene, this was counted twice. B) A bar chart showing the proportion of isolates per year that had different H-types. The colours are the same as those represented in Figure 3C.

We performed long-read sequencing for 14 isolates which either had no O-type call or had multiple O-type calls, to determine the genomic region between *galF* and *gnd* where the O-antigen type locus is usually found in *E. coli* (Figure 4). Having no O-antigen is highly unusual and leads to increased susceptibility to antimicrobial stress, and thus seems unlikely to be present in clinical isolates. Ten different O-antigen loci were revealed in these 14 isolates, with four of them represented by two isolates each. Two isolates were confirmed as O178 and one appeared to be a hybrid of O8 and O160, whilst seven of these ten O-types were so far undescribed (Figure 4) and one showed similarity to the OX-13 gene in *Salmonella* (BKRHXR). Three of the isolates (CAAH3Y, CNS75S and BKQ37E) have loci heavily disrupted by insertion elements and seem to lack the components for an export machinery (*wzm*/*wzt* or *wzx*/*wzy*). It remains to be investigated whether they acquired an entirely unrelated O-antigen locus from a different organism that integrated into a different part of the genome and encodes for export machineries sufficiently different to not even get recognized by read-based searches, or whether these isolates indeed do not encode for a classical O-antigen. One of the isolates (CAAXI3) has a similarly disrupted *galF*/*gnd* site but encodes for a potentially functional O-antigen locus on a plasmid (CAAXI3_2), raising interesting questions regarding the expression of this O-antigen locus and whether this will remain plasmid-located or eventually become integrated into the disrupted chromosomal location.

**Figure 4.**
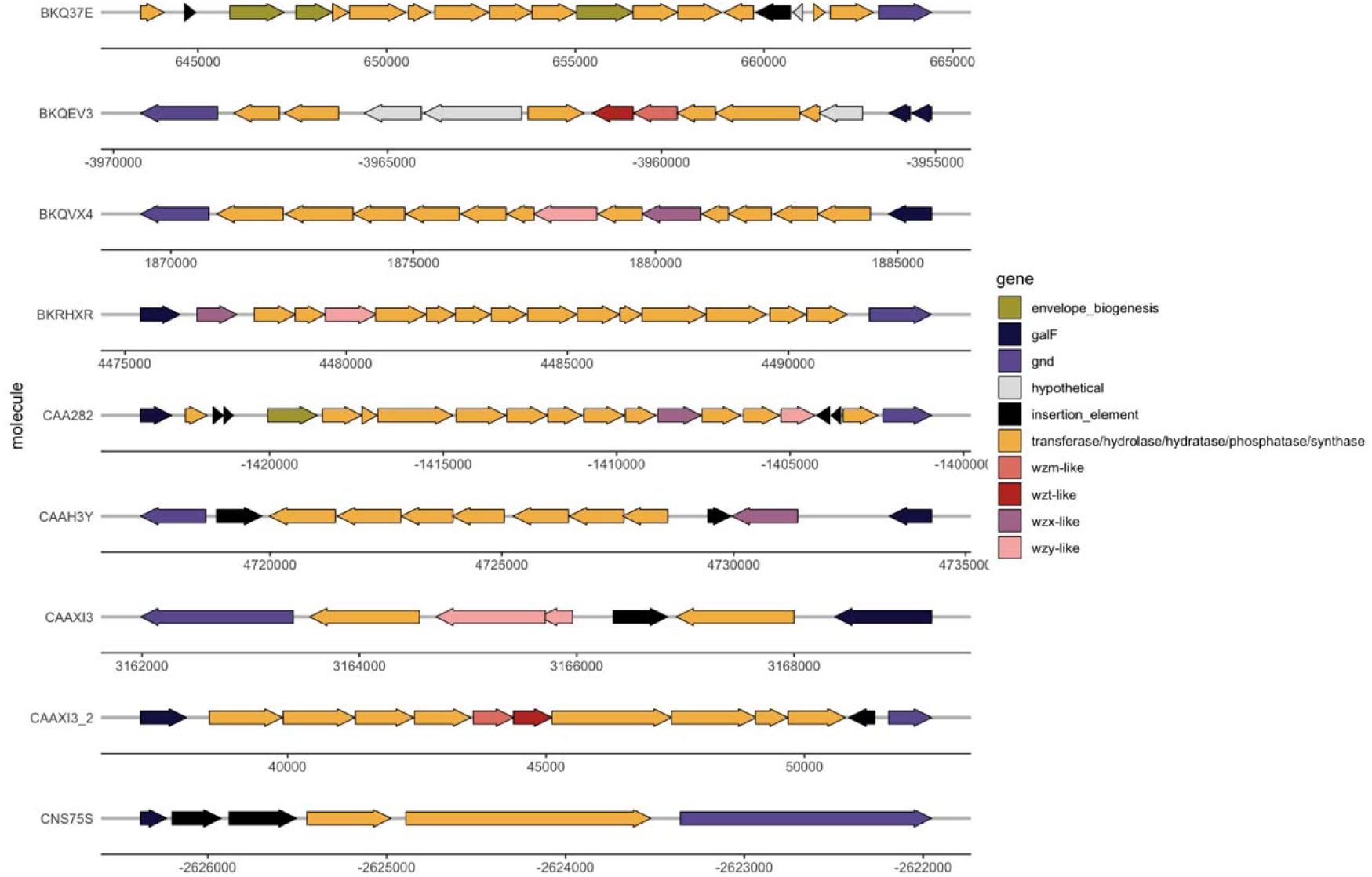
A schematic showing the operon structure of the novel O-antigen genes. Each row represents a different novel O-type from a single isolate. For isolates with > 0.9 sequence homology for the O-antigen gene, only one isolate was selected for representation.

There were 34 H-types found in the collection, of which 4 H-types were each identified in more than 10% of isolates. These were H4 with 29/173 (16.8%), H18 with 24/173 (13.9%), H5 with 18/173 (10.4%) isolates and H9 with 17/173 (9.8%) isolates (Figure 3C). Five H-types were found in at least 75% of the years (6 out of 7 or more) that had more than 10 isolates (H4, H18, H5, H9 and H7). H4 and H18 were the only H-types responsible for greater than 20% of the isolates in any year with more than 10 isolates (in two years each) but no H-type was identified in over 50% of isolates in a single year. The largest proportion of isolates belonging to a single H-type in a single year was H4 with 5/14 (35.7%) of cases in 2013 (Figure 3D). There were 4/169 (2.4%) isolates which had more than one H-type and may be able to undergo phase variation for immune escape, hence the denominator of 173 above.

Considering the O and H types in light of body site of isolation (e.g. bloodstream or CSF), 20 different O-types were encoded on isolates derived from CSF. O8 was found in 8/33 (24.2%) isolates from CSF, compared to 4/133 (3.0%) of bloodstream isolates (χ^2^ *p* = 0.0001). This was partly due to ST410 (six isolates) which all encoded for O8, however there were also two other O8 isolates (one ST58 and one ST155) that occurred in CSF. There were six other O-types that occurred in two CSF samples (O78, OX13-like, O25B, O1A, O18A1 and O15) with the rest occurring in only one CSF sample. Of these, only OX13-like was found in a significantly different percent of CSF samples compared to bloodstream isolates (2/33 [6%] vs 0/133 [0%], χ^2^ *p* = 0.049). There were 17 H-types isolated from CSF. H9 was found in 8/33 (24.2%) isolates from CSF, compared to 8/133 (6.0%) of bloodstream isolates (χ^2^ *p* = 0.004). This was again partly due to the six ST410 isolates, however there were also two ST23 isolates that expressed H9 found in CSF. H5, H4, H7, H18 and H1 all occurred in more than one CSF isolate, with the other H-types occurring in just one CSF isolate.

Excluding STs for which there was just one isolate, we examined whether multiple O-types or H-types were found in isolates of a single ST. The median number of O-types per ST for the STs that met this criterion was 2, with 10/23 (43%) encoding for just a single O-type and 13/23 (57%) encoding for multiple O-types. The median number of H-types per ST for the STs that met our criteria was 1 with 14/26 (54%) encoding for just a single H-type and 12/26 (46%) encoding for multiple H-types. Three of the most frequently occurring STs encoded for multiple O-types and H-types. ST10 showed the highest diversity of O-types and H-types, with 10 different O-types and 7 different H-types. ST131 covered 3 different O-types (O25B, O11 and O16) and 2 different H-types (H4 and H5), which occurred from 2013 to 2020 and often with multiple O-types or H-types in the same year. ST69 isolates included 4 different O-types and 2 different H-types. ST410 on the other hand occurred frequently from 2016 to 2020 but all isolates encoded for only a single O-type (08) and a single H-type (H9).

The EXPEC9V conjugate vaccine (which covers the O1A, O2, O4, O6A, O15, O16, O18A, O25B and O75) might be expected to confer immunity to up to 57/169 (33.7%) of these cases, the original 10V composition (including O8) would have covered up to 77/169 (45.6%), demonstrating a loss of 11.9% by the removal of just one O-antigen of high prevalence in our setting (Figure 5A and Supplementary Figure 2). The EXPEC4V vaccination (which covers O1A, O2, O6A, and O25B) would cover at maximum 29/169 (17.2%) of cases (assuming the O6 isolates are of the O6A subtype, something that we have not been able to confirm; Figure 5A; 5B; methods). Analyzing the data by year (including only years with greater than 10 isolates) the EXPEC9V vaccine covered fewer than 50% of the isolates’ O-types in every year, and 3 out of 7 years covered less than 30% of the vaccine O-types, with the lowest coverage in 2013 where only 23% of the isolates were from vaccine O-types, and importantly we observe no major change in coverage over time (Figure 5C). The EXPEC4V vaccine covered less than 30% of the isolates’ O-types in every year, and 5 out of 7 years covered less than 20% of the vaccine O-types, with the lowest coverage in 2017 where only 5% of the isolates were from vaccine O-types, with fluctuations over time that do not indicate any improvement in coverage in future (Figure 5C). Regarding the isolates which were resistant to first- and second line antimicrobial therapy (benzylpenicillin, gentamicin and ceftriaxone), the EXPEC9V vaccine would be expected to confer immunity to 13/34 (38.2%) of these cases and the EXPEC4V vaccine would be expected to confer immunity to 9/34 (26.5%) of these isolates.

**Figure 5A).**
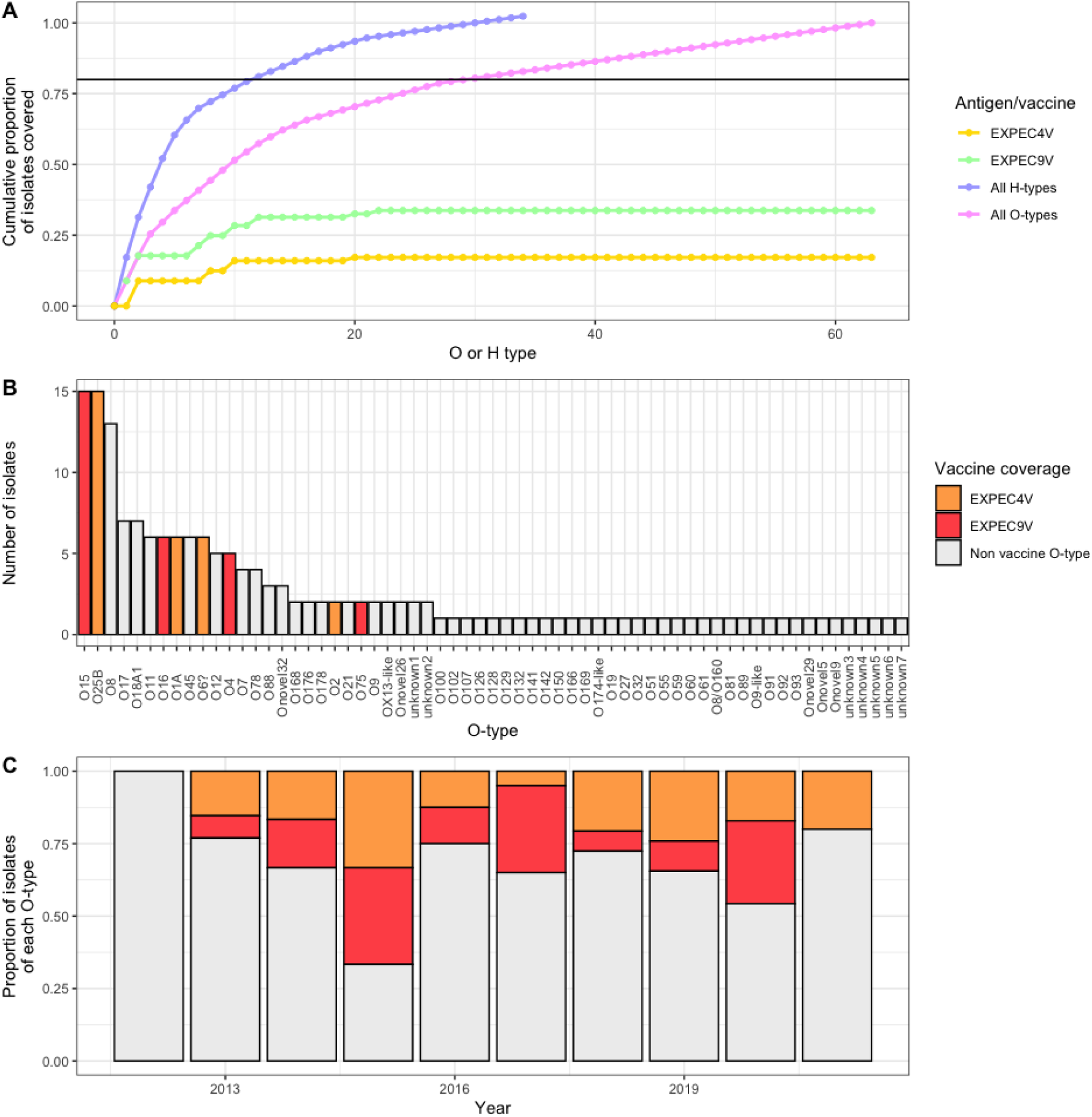
Rarefaction curve showing the theoretical protection given against vaccines covering the most frequently isolated H-types and O-types, as well as the potential protection given by the EXPEC9V and EXPEC4V. The horizontal line shows the point at which 80% of isolates would be covered. For isolates with more than one H-type both were counted, there were multiple isolates with more than one H-type so the line for H-type goes above 1. Supplementary Figure 3 shows the same graph but where isolates had more than one H-type called they were only counted once. B) A bar chart showing the frequency of the different O-types. C) A bar chart showing the proportion of isolates per year that had different O-types. The colours are the same as those represented in Figure 5B.

To cover 80% of cases, an O-antigen based vaccine would need to offer protection against the top 30 O-types. In contrast, to cover the top 80% of these cases, an H-type vaccine would only need to cover the top 12 H-types (Figure 5A). If the four most frequently occurring O-types in our setting were selected from our cohort for a vaccine (O15, O25B, O8 and O17) this vaccine would cover 50/169 (29.6%) of cases, ranging from 21% to 40% per year, and the nine most frequently occurring O-types (O15, O25B, O8, O17, O18A1, O11, O16, O1A and O45) would represent just under half of the isolates 81/169 (47.9%), ranging from 33% to 67% per year, with numbers fluctuating over our study period showing no indication that there would be an increase in coverage in future.

### Antimicrobial resistance and plasmid replicons

At the time of the study the first line treatment for neonatal sepsis and meningitis in QECH was benzylpenicillin and gentamicin, with second line treatment ceftriaxone. *E. coli* is intrinsically resistant to benzylpenicillin and isolates with acquired resistance to gentamicin and ceftriaxone were therefore difficult to treat (42/194 [21.6%]). There was occasional but limited use of amikacin or meropenem for neonates with proven or high suspicion of ceftriaxone resistance or who were very unwell. The use of meropenem and amikacin increased over the study period.

We identified AMR genes against all major classes of antibiotics and several efflux pump systems, in line with the phenotypic resistances detected. The number of AMR genes varied by ST, with ST410 (mean 24.0, SD 1.9) and ST131 (mean 20.6, SD 4.8) having the greatest number of average AMR genes per isolate. ST10 had a lower number of AMR genes per isolate (mean 8.9, SD 1.6), whilst ST69 was intermediate (mean 12.9, SD 1.5). ST410 was present only from 2016 onwards which may partly explain the higher number of resistance genes, whilst the other STs, including ST131 were present throughout the study period.

The number of *E. coli* isolates that were resistant to ampicillin was 143/190 (75.3%) and was stable over the period (-0.3% change per year; *p* = 0.96; Figure 6A). *bla*_EC_ genes, which are chromosomally encoded in *E. coli,* were found in all 169 isolates (*bla*EC-15, *bla*_EC-5_, *bla*_EC-18_ and *bla*_EC-8_). The most frequently occurring plasmid-encoded beta-lactamase penicillinase genes included *bla*_TEM-1_ found in 121/169 (71.6%) isolates, and *bla*_OXA-1_ found in 21/169 (12.4%) of isolates (Supplementary Figure 3). Whilst only 49/197 (24.9%) were resistant to gentamicin, this however showed a temporal trend, increasing from 3/15 (20%) in 2013 to 16/39 (41%) in 2020 (22.8% change per year; *p* = 0.0035; Figure 6A). Gentamicin resistance mechanisms were mainly variants of the gene *aac*(3), *aac*(3)-IId found in 23/169 (13.6%) isolates and the *aac*(3)-IIe gene found in 12/169 (7.1%) isolates (Supplementary Figure 3).

**Figure 6.**
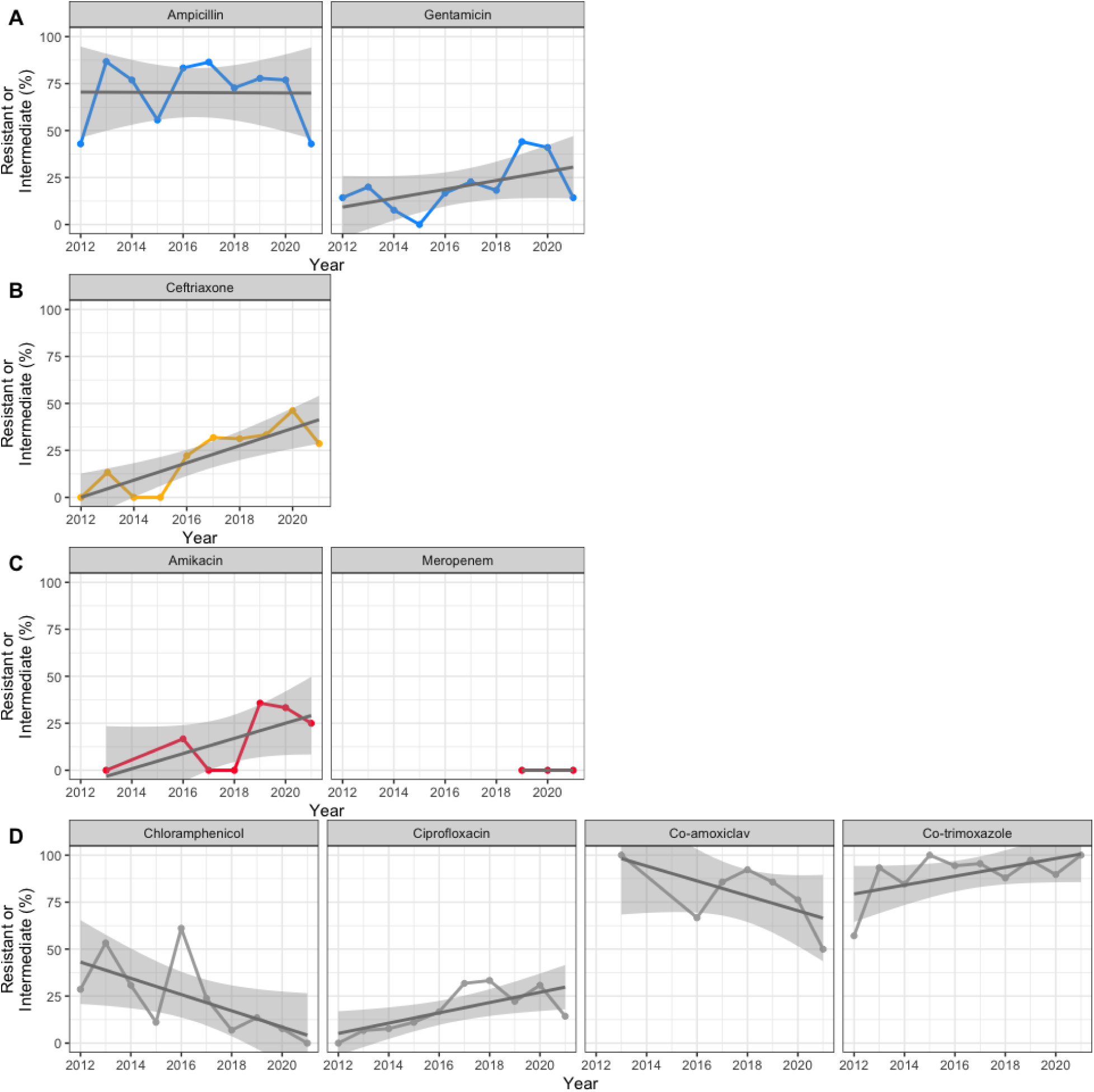
The proportion of E. coli isolates that were phenotypically resistant to different antibiotics by year. A) First-line antibiotics for neonatal infection B) Second-line antibiotics for neonatal infection C) Occasionally used antibiotics for neonatal infection D) Antibiotics not used in neonates, but used elsewhere in the hospital or in the community.

Ceftriaxone was the second-line treatment at the time of study, and 55/198 (27.8%) were resistant to ceftriaxone and increased over the period from 0/7 (0%) in 2012 to 18/39 (46.2%) in 2020 (31.8% change per year; *p* = 0.00014; Figure 6B); and 42/55 (76.4%) of the isolates resistant to ceftriaxone were also resistant to ampicillin and gentamicin, hampering the effectiveness of all first- and second-line treatments. The increase in ceftriaxone resistance was due to the widespread ESBL gene *bla*_CTX-M-15_, which was detected in 39/169 (23.0%) isolates (Supplementary Figure 4), whilst other alleles, *bla*_CTX-M-14_ and *bla*_CTX-M-27_, could be identified in only 1/169 (0.6%) isolate each, both from 2018 (Supplementary Figure 2). ESBL genes were frequent in ST410 (8/8 [100%]) and ST131 (9/18 [50%]) as observed in other studies^55, 56^, and infrequent in ST69 (3/20 [15%]) and ST10 isolates (1/15 [7%]). The proportion of isolates from late onset infection resistant to ceftriaxone was 16.9% (95% CI 3.5 - 30.2%) higher than from early onset cases (37.2% vs 20.4%, *p* = 0.012); the same pattern was observed for gentamicin with 15.8% (95% CI 2.8 - 28.7%) more resistant isolates in late onset infections compared to early onset cases (33.6% vs 17.9%, *p* = 0.016), whereas similar proportions were observed for ampicillin with 78.5% vs 73.2% in late and early, respectively (*p* = 0.50).

Alternatives for isolates resistant against all of the above antimicrobials are amikacin or carbapenems, and so far only a small proportion, 7/67 (10.4%), were resistant to amikacin. This increased from 1/6 (17%) in 2016 (amikacin was not routinely tested until 2016) to 7/21 (33%) in 2019 (52.7% change per year; *p* = 0.042; Figure 6C). The main gene identified was the amikacin resistance gene *aac*(6’)-Ib-cr5 in 21/170 (12.4%) isolates (Supplementary Figure 2). Of the 42 isolates resistant to all first and second-line agents 11/42 (26.2%) were causing meningitis and a further 5/42 (11.9%) were resistant to amikacin, leaving only meropenem as effective treatment for these (amikacin does not reliably penetrate the blood-brain barrier). In line with the low levels of carbapenem resistance identified in other studies from this setting, no isolates showed phenotypic meropenem resistance.

Other antimicrobials are not regularly used on the neonatal unit but are tested for routinely for *E. coli*. Fluoroquinolones are still widely used in other wards, and 45/199 (22.6%) were resistant to ciprofloxacin which increased over the period from 0/7 (0%) in 2012 to 12/39 (30.8%) in 2020 (20% change per year; *p* = 0.013; Figure 6D). There were several isolates with fluoroquinolone resistance mutations, the most frequent were the *gyr*A mutations *gyr*A_S38L found in 38/169 (22.5%) isolates and *gyr*A_D87N found in 24/169 (14.2%) isolates, the *par*C mutation *par*C_S80I found in 26/169 (15.4%) (Supplementary Figure 2). We further note 37/169 (21.9%) isolates with *parE* mutations which are not described as sufficient to provide resistance for *E. coli,* but could lead to reduced susceptibility or higher resistance levels if an additional mutation is present. All ST410 isolates encoded four different, acquired fluoroquinolone resistance genes (two *gyr*A mutations, one *par*C and one *par*E each). Likewise, we identified at least one acquired fluoroquinolone resistance gene in all ST131 isolates *gyr*A, *par*C and *par*E). Fluoroquinolone resistance genes (*gyr*A, *par*C and *qnr*S1) were found in 8/20 (40%) of ST69 isolates and 3/15 (20%) of ST10 isolates.

Chloramphenicol resistance has been observed in other isolates in this setting to decrease, and in line with this we observed 41/195 (21%) isolates resistant to chloramphenicol with a decreasing trend, from 2/7 (28.6%) in 2012 to 3/39 (7.7%) in 2020 (-30.5% change per year; *p* < 0.0001; Figure 6D). The most frequently occurring chloramphenicol genes were *cat*A1 found in 22/170 (12.9%) isolates and *cat*B3, found in 20/170 (11.8%) isolates (Supplementary Figure 2).

Overall 54/67 (80.6%) were resistant to co-amoxiclav (not used on the neonatal unit and infrequently used in other hospital wards) and the proportion decreased slightly over the period (-20.7% change per year; *p* = 0.33; Figure 6D). Resistance to co-trimoxazole (used as prophylaxis against Pneumocystis pneumonia in HIV patients) was high over the period at 183/200 (91.5%) and increased slightly (14.9% change per year; p = 0.13; Supplementary Figure 2). Colistin resistance was not tested in our cohort, but genes conferring colistin resistance were found relatively frequently, with *pmr*B_E123D found in 48/169 (28.4%) isolates and *pmr*B_Y358N found in 36/169 (21.3%) isolates (Supplementary Figure 2).

There were 34 different plasmid replicons found in the dataset. The most frequently identified were the commonly detected IncF plasmid replicons that frequently carry resistance cassettes, with IncFIB_AP001918 found in 107/169 (63.3%) of isolates, IncFI found in 91/169 (53.8%) of isolates and IncFII_p found in 46/169 (27.2%) of isolates (Supplementary Table 1). Multiple different plasmid replicons were also found of the Col, IncH and IncX types, with other types found less frequently.

## Discussion

This study highlights the challenges of controlling neonatal sepsis with vaccines in a low income setting when there is a paucity of data from this region describing the nature and diversity of isolates causing disease. We present the trends in predicted serogroup epidemiology of a collection of *E. coli* isolated from neonates in a single, large teaching hospital in Malawi from 2012 to 2021. Our study reveals that O-antigen vaccines would need a high valency (30 O-types) to achieve protection against greater than 80% of isolates, and vaccines in current development for use in elderly populations in high-income countries would offer protection against only one third of the *E. coli* isolated in this study.

Our study, consistent with similar studies from high income countries^57, 58^, found approximately 50% of *E. coli* cases to be from early-onset sepsis (EoS) and 50% from late-onset sepsis (LoS). We found higher rates of ceftriaxone and gentamicin resistance in the LoS cases compared to the EoS cases (though this finding is not typical of other studies^59^,^60^), which might imply that in our setting these two groups have different epidemiology (e.g. EoS cases being maternally transmitted and LoS cases deriving from the hospital environment or from infection in the community). Almost a fifth of our isolates were from neonatal meningitis cases, and *E. coli* is an important cause of meningitis in neonates, including in low-income countries^61^. Neonatal meningitis is particularly concerning as it is associated with greater morbidity and mortality, requires longer treatment (minimum 21 days of antimicrobial therapy for *E. coli* meningitis compared to 7 days for bloodstream infection) and certain drugs such as amikacin cannot be used for meningitis due to probable poor blood-brain barrier penetration, leaving very few options for isolates resistant against third-generation cephalosporins.

The numbers of *E. coli* cases per 1,000 blood culture or CSF tests in all age groups decreased from the period 2000 to 2012 but was relatively stable from 2012 onwards (which is the time period for which we had genomic data). There were peaks in the absolute numbers of cases in 2005 and 2019, although the peak in 2019 appears to be primarily related to greater patient numbers as there was no increase in the number of cases per 1,000 blood culture or CSF tests done. This contrasts with *K. pneumoniae* at the same site over the same period^62^ which showed a large peak in numbers in 2019.

We saw very high ST diversity, with many STs occurring only once and eleven STs that were previously unknown. This high ST diversity which was consistent over the whole study period may indicate that neonates are exposed to diverse sources of *E. coli.* It also illustrates the need for more studies of *E. coli* diversity from sub-Saharan Africa. Although the STs with highest numbers in our study are part of globally prevalent high-risk clones^63^ (ST131, ST10, ST69 and ST410), no ST was persistently present as a major lineage in our study. Our findings are comparable to previous findings from the same site in Malawi over a longer period^64^ and another study from Lilongwe^65^ which also found these STs to be common. ST410 has been associated with increased mortality in the Malawian context as one study found that all 8/8 (100%) of patients in the study that had bloodstream infection with ST410 died^55^ vs 135/326 (41%) of the overall cohort. In our study ST410 was associated with meningitis, the ESBL genes *bla*_CTX-M-15_ and fluoroquinolone resistance genes, leaving only meropenem as a treatment option.

The vaccines against *E. coli* which are in clinical trial stages target O-antigens. Whilst being developed to protect against UTIs and urosepsis in high-income settings, these or similar O-antigen based vaccines could feasibly be administered to mothers to prevent neonatal sepsis. The choice of O-antigen glyco-conjugate vaccines is based on the knowledge that O-antigens are the major cell surface component of *E. coli*^66^ and appear to be essential for *E. coli* survival in human serum^67.^ There are also multiple other glyco-conjugate vaccines have been successful (including *Haemophilus influenzae* type b^68^, *Streptococcus pneumoniae*^69^ and *Neisseria meningitidis*^70^, though these are all based on bacterial capsule rather than O-antigen). Our study indicated high O-type and H-type diversity, with large flux of both, and no O-type or H-type representing the majority of cases in any year. As is the case in other collections^31^ there was higher diversity of O-type than H-type, meaning vaccine approaches targeting O-type require a much higher valency than those targeting H-types to protect against a similar proportion of isolates (30 O-types vs 12 H-types to protect against 80% of isolates). There were also 10 previously undescribed O-types and no undescribed H-types in our cohort. This higher diversity (including some that is previously undescribed) may call into question the practicality of using an O-type based vaccine approach in our patient population and may direct efforts towards exploring vaccines based on other antigens.

The EXPEC9V vaccine was designed for an elderly population in high-income countries, and for this role, it is likely to be effective. It was initially designed as a 10-valent vaccine; however, the serotype O8 was removed after the functional antibody assay did not work^13, 14, 15^. In our cohort, this change would have had a significant effect on the utility of such a vaccine (dropping the proportion of isolates protected against from 45.6% to 33.7%). O8 was the third most frequently occurring O-type in our cohort, it was enriched amongst our meningitis cases and was present in all our ST410 isolates (which were highly AMR) and thus seems to be particularly common in high consequence infections. Its removal would therefore drastically reduce the likely impact of this vaccine in our setting. Other studies in the target population for this vaccine (elderly adults in high-income countries) have shown good protection against invasive cases (64.7%^71^ - 67.5%^72^). The O-types in this vaccine appear wholly appropriate for this patient population. Interestingly, one of these studies^71^, though it was conducted across three continents and seven countries reported lower O-type diversity than in our collection (49 O-types; 47 identified O-types as well as two unknown O-types compared to the 63 distinct O-types found in our collection). This might reflect differences in the target setting as well as the patient population. Most appropriate to compare to our study is a study on a cohort of paediatric (most cases were from neonates) *E. coli* meningitis cases from France, where O1, O18, O45 and O7 were the most common O-types^73^ Amongst these, O18 was the only O-type found frequently in our study whilst other O-types such as O17, O12 and O11 were frequently found in our study but not in that one.

We also identified nine potentially novel O-types and one combined O-type that were not identified by the EcOH database or ECTyper, highlighting our incomplete overview of O-antigen diversity, and that we lack knowledge of how frequently new types emerge. These putative novel O-types furthermore emphasize the need to perform sero-typing and WGS on more isolates from sub-Saharan Africa as there might be substantial undescribed diversity. One of the unknown O-types was similar to the OX-13 antigen on *Salmonella enterica*. There are some O-types that are known to be shared by both *S. enterica* and *E. coli* (*E. coli* O-types O55, O111 and O157^74, 75^). This is likely another O-type that is shared by both bacterial species. One O-type appeared to have sections from both the O8 and O160 O-type sugar molecules, it is not clear whether this is a hybrid of the two sugar molecules or whether this organism would be able to express both molecules.

Phenotypic AMR increased for several different antibiotics over the study period (ceftriaxone, co-trimoxazole, gentamicin, ciprofloxacin and amikacin, though co-trimoxazole did not have a *p* value < 0.05) and was explained by a number of different genomic mechanisms. Ceftriaxone had been introduced in QECH as standard therapy for many infections in 2004, so this introduction alone cannot be responsible for the increase in resistance seen here. This is a global trend and whilst rates of AMR for *E. coli* are lower than for *Klebsiella pneumoniae* at the same site^4,62^, they are of significant concern. We identified no isolates with carbapenem resistance, which is not surprising as these genes are not widespread in Malawi, however a previous study has identified a single carbapenemase carrying *E. coli*^74^. Chloramphenicol and co-amoxiclav resistance decreased over the study period (though only chloramphenicol had a *p* < 0.05), this agent is however contraindicated in neonates and thus does not provide a treatment alternative. The increase in AMR, particularly in a setting where watch and reserve antibiotics are often unavailable due to cost, highlights the importance of prevention of neonatal infection in the first place with strategies such as vaccines or investment in improvements in infection, prevention and control (IPC).

In conclusion, the ongoing burden of neonatal sepsis combined with worsening AMR in *E. coli* motivates the development of *E. coli* vaccines for this population. However, the prevalent O-types in this collection from sub-Saharan Africa are highly diverse, partially novel, and different to those currently covered by vaccines in clinical trials. If maternally-administered vaccines are to be developed, they need to be based on robust genomic surveillance of prevalent antigens and temporal trends in this population, and far more data from sub-Saharan Africa is required to ensure equity of coverage compared to vaccines developed for HICs. Development of a suitable vaccine will be a lengthy process, and until a successful product is available other methods of preventing mortality from neonatal sepsis such as IPC strategy and early recognition of neonatal infection should be urgently supported.

## Supplementary materials

**Supplementary Figure 1.**
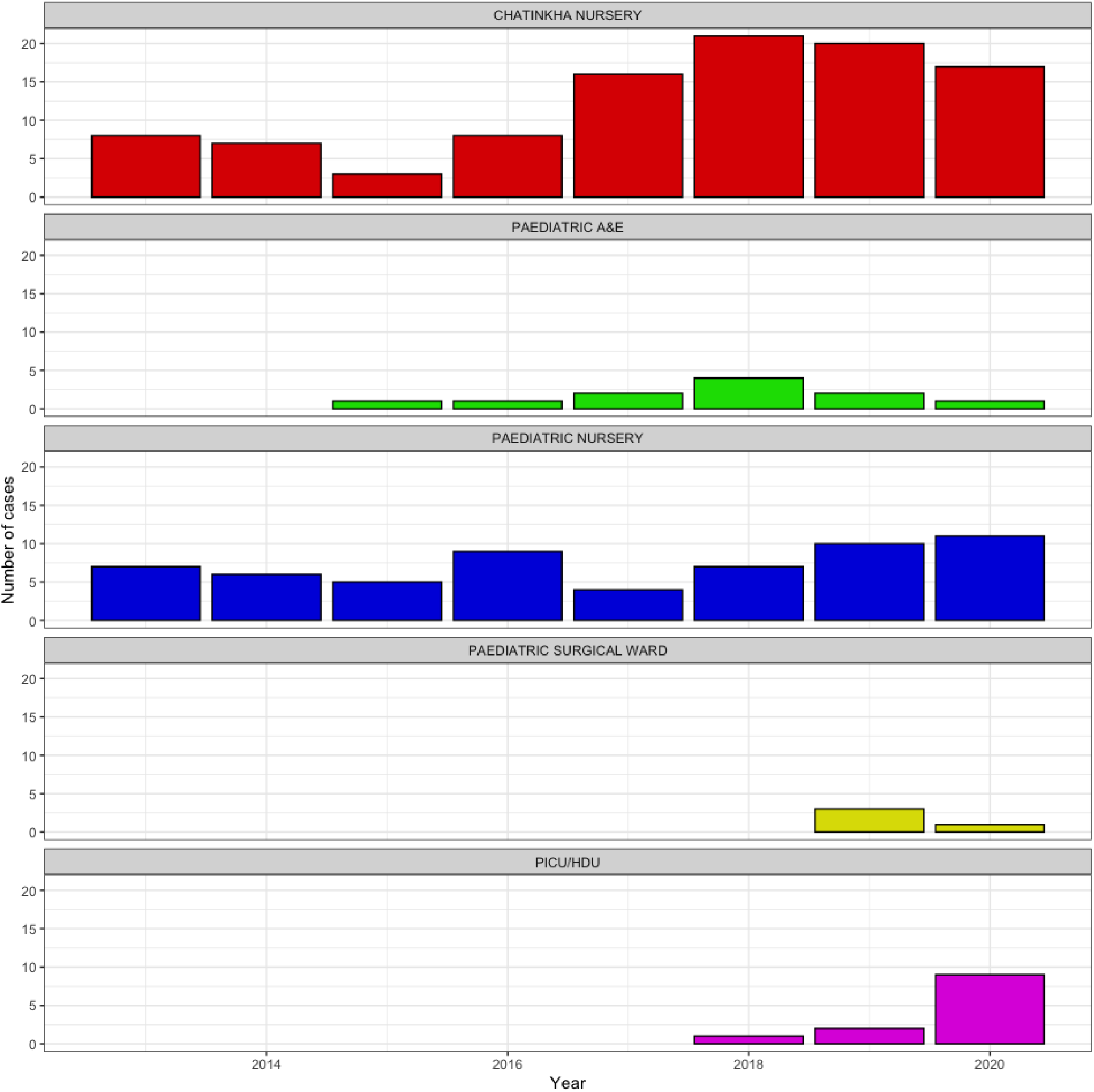
E. coli cases per year by ward.

**Supplementary Figure 2).**
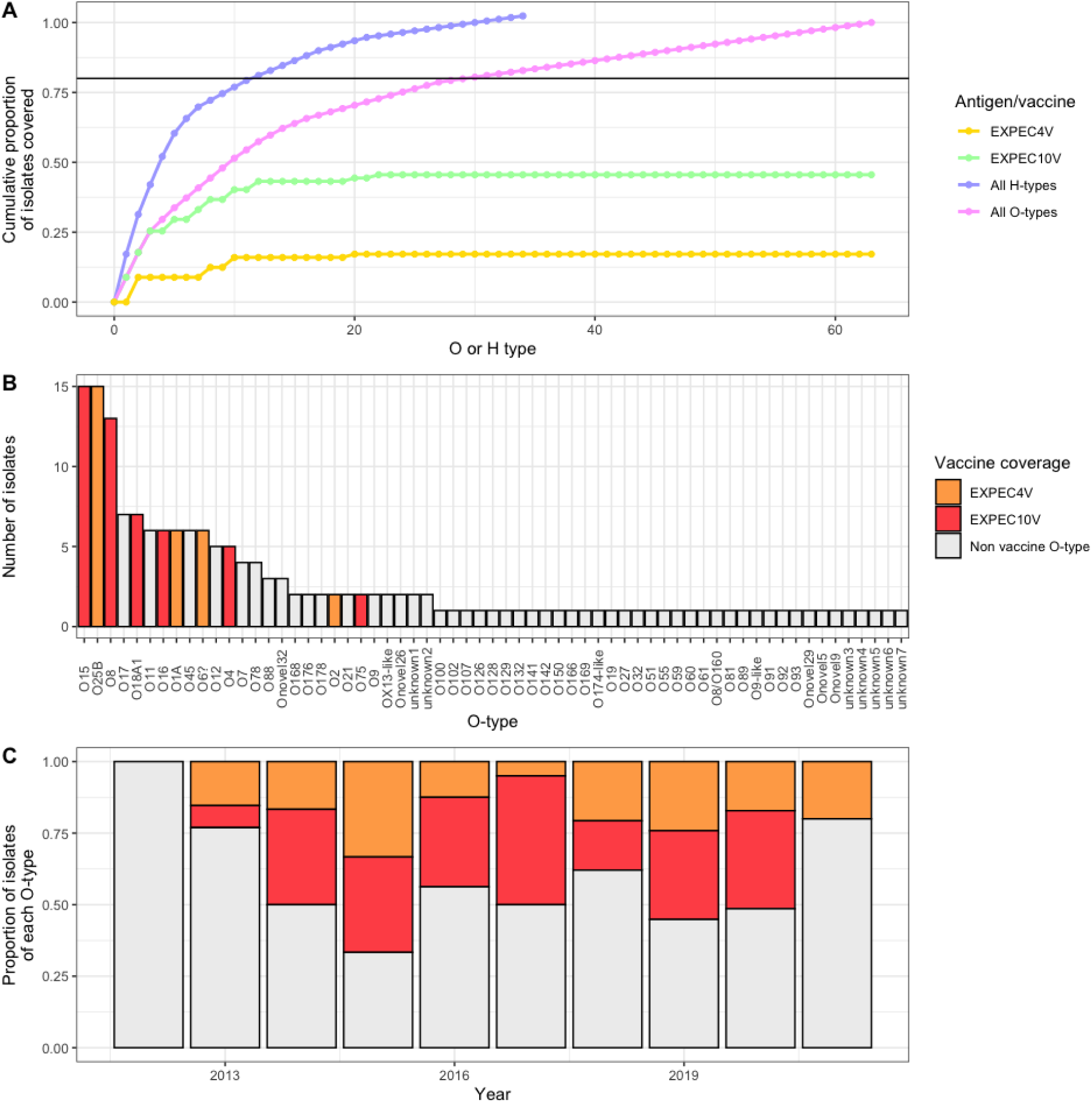
Rarefaction curve showing the theoretical protection given against vaccines covering the most frequently isolated H-types and O-types, as well as the potential protection given by the EXPEC10V and EXPEC4V. The horizontal line shows the point at which 80% of isolates would be covered. For isolates with more than one H-type both were counted, there were multiple isolates with more than one H-type so the line for H-type goes above 1. B) A bar chart showing the frequency of the different O-types. C) A bar chart showing the proportion of isolates per year that had different O-types. The colours are the same as those represented in Figure 5B.

**Supplementary Figure 3.**
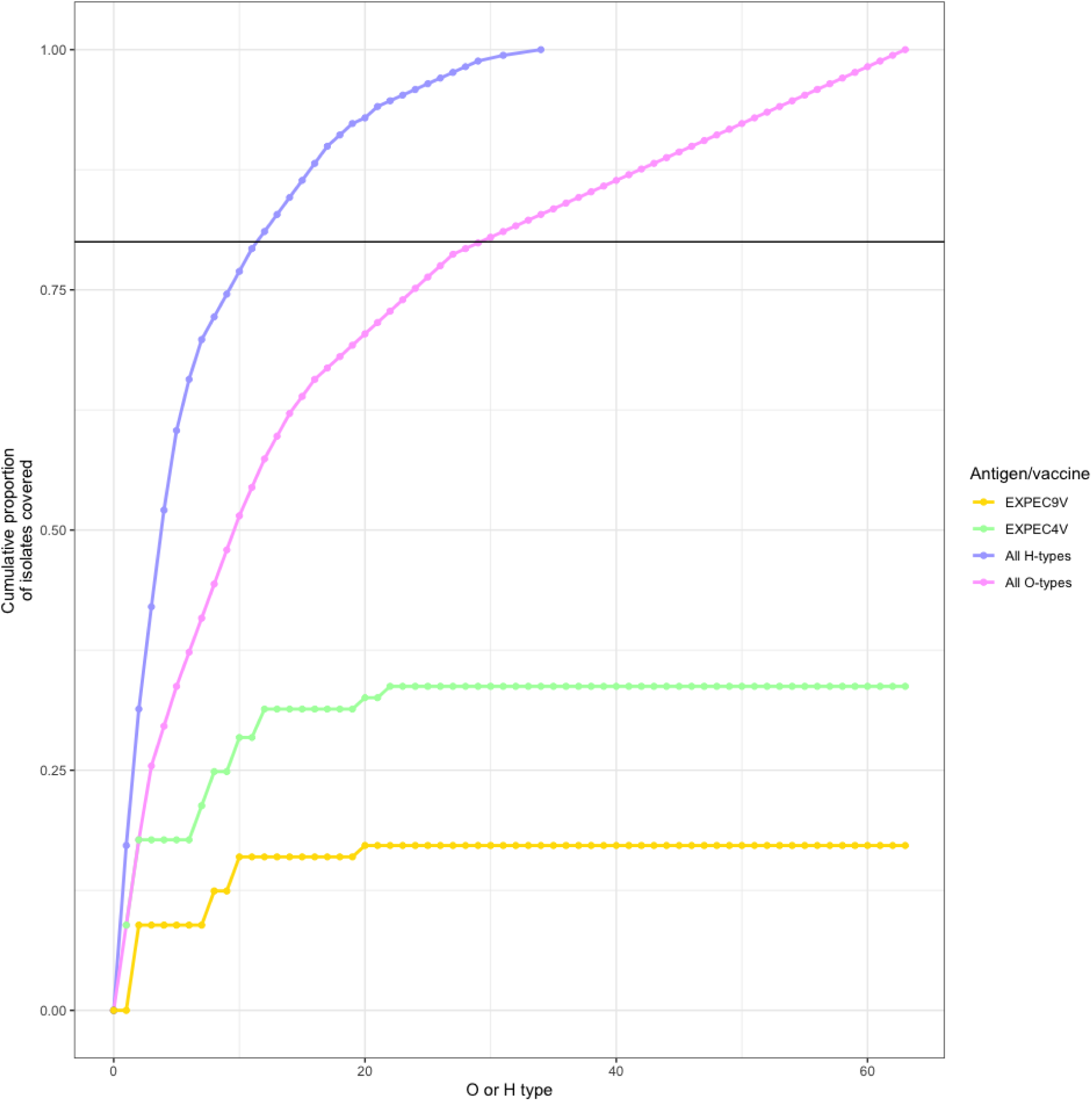
Rarefaction curve showing the theoretical protection given against vaccines covering the most frequently isolated H-types and O-types, as well as the potential protection given by the EXPEC9V and EXPEC4V. The horizontal line shows the point at which 80% of isolates would be covered. For isolates with more than one H-type only the most frequently occurring O-type or H-type was counted.

**Supplementary Figure 3.**
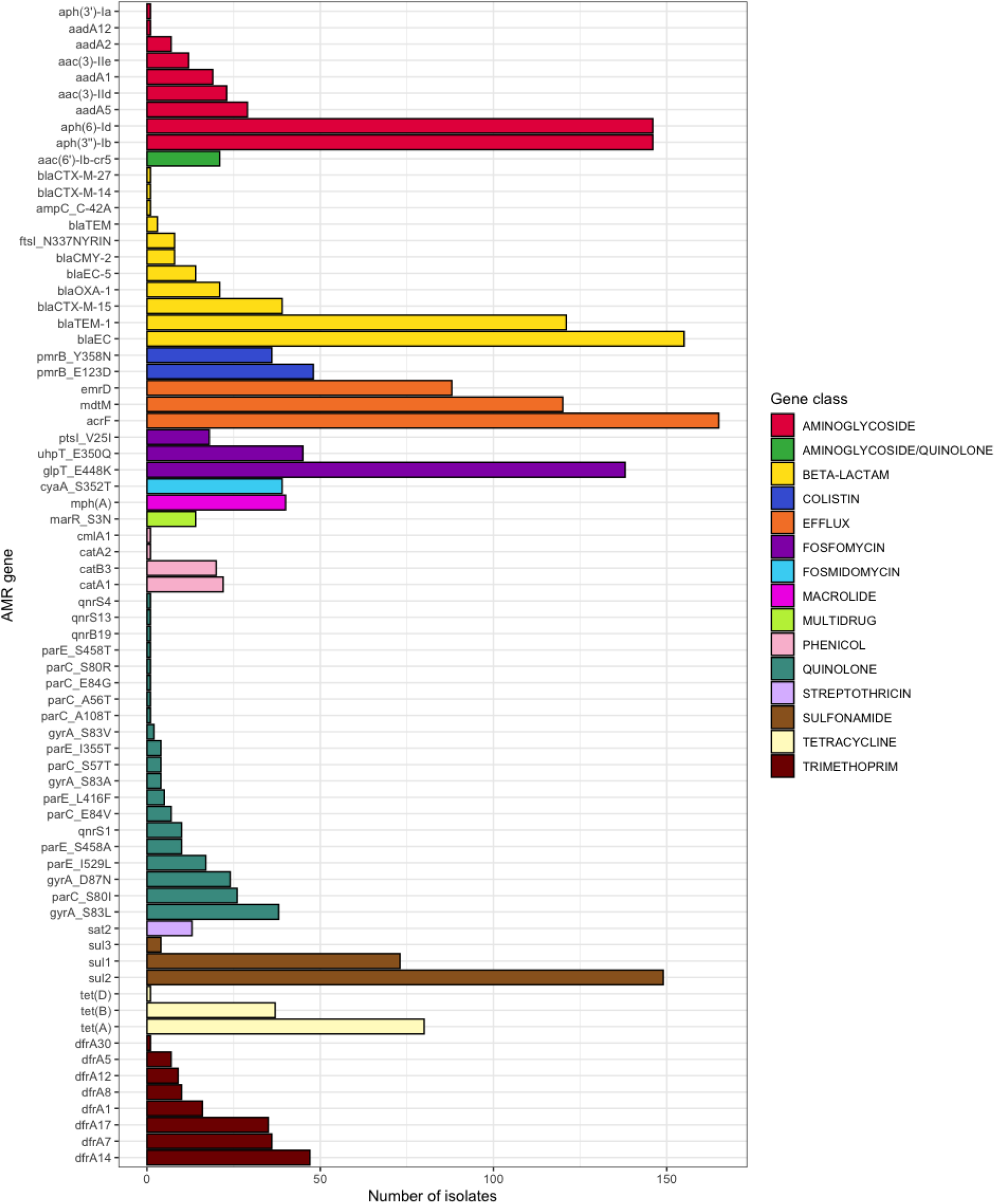
Frequency of antimicrobial resistance (AMR) genes in the collection separated by class.

## Author contributions

The study was conceived by NAF and OP. NAF, NRT and EH were responsible for funding acquisition. Investigation and methodology development was carried out by AZ, ET, PS, AJF, JC, PM and EH. Data curation and project administration was carried out by OP and EH. Resources were managed by AZ, KK and OP. Formal analysis, validation and visualization were carried out by OP, AJF and EH. Supervision was carried out by NAF and EH. The writing of the original draft of the manuscript was by OP, NAF and EH. It was edited and revised by all authors. All authors read and agreed on the final manuscript.

## Data Availability

All sequencing data is freely available under the sequencing project IDs ERP120687 (short read data; accessions in Supplementary Table 1) and PRJNA1121524 (long-read data; accessions in Supplementary Table 2), detailed per-isolate information is provided in Supplementary Table 3. Blood culture and CSF data used to show the trends and numbers of E. coli cases per year is available in Supplementary Table 4.

## Acknowledgements

We would like to acknowledge the clinical team at QECH and the Mercy James hospital for caring for the babies who were affected by *E. coli* in this study. We would also like to acknowledge the MLW microbiology laboratory team, who isolated and identified the *E. coli*. We would also like to acknowledge the Pathogen Informatics teams at the Wellcome Sanger Institute for their expert support.

